# Spatial transcriptomics maps distinct signatures of human intermuscular adipose expansion in mice

**DOI:** 10.64898/2026.04.16.26351017

**Authors:** Ekta Pathak, Robby Z. Tom, Myeongseop Kim, Stephan Sachs, Yao Zhang, Marc Walter, Paul T. Pfluger, Annette Feuchtinger, Ken A. Dyar, Bryan C. Bergman, Miguel A. Pleitez, Dominik Lutter, Susanna M. Hofmann

## Abstract

Intermuscular adipose tissue expansion is closely associated with cardiometabolic disease, yet its cellular organization and regulatory mechanisms remain poorly defined. Using bulk transcriptomics on human intermuscular adipose tissue, we identified a distinct gene signature and functional regulators including adipogenic transcription factor early B-cell factor 2 (EBF2). By mapping this human signature to the spatial transcriptome of intermuscular adipose tissue from mice with cardiometabolic disease, we unraveled discrete stromal niches surrounding muscle fibers, characterized by intermuscular adipose tissue expansion and the coordinated activation of adipogenic, extracellular matrix, inflammatory, and metabolic pathways. Spatial analyses showed that fibro-adipogenic progenitor abundance does not predict adipocyte formation, supporting a model of localized and context-dependent lineage transitions. Cross-species comparison revealed partial conservation of human intermuscular adipose tissue gene programs, validating the mouse model and highlighting species-specific features. Functional experiments in human primary myoblasts showed that EBF2 is sufficient to induce adipogenic reprogramming. Our findings establish intermuscular adipose tissue as an active, spatially organized remodeling niche and identify lineage plasticity as a central mechanism driving its expansion in metabolic disease.

## Introduction

Skeletal muscle is a metabolically active organ central to whole-body energy homeostasis. Beyond its contractile role, skeletal muscle functions as an endocrine and metabolic tissue that influences systemic glucose and lipid regulation. Within this complex architecture, intermuscular adipose tissue (IMAT), defined as adipocytes located between muscle fascicles has emerged as a metabolically relevant fat depot (Goodpaster et al., 2023). Increased IMAT accumulation is observed in obesity, aging, and insulin-resistant states and is strongly associated with reduced muscle function, decreased contractile performance, and increased cardiometabolic risk, independent of overall adiposity (Goodpaster et al., 2023; Biltz et al., 2020; Camacho-Cardenosa et al., 2025). Recent advances in imaging and molecular profiling have revealed that IMAT is not just a lipid storage, but an active endocrine and paracrine tissue capable of influencing muscle metabolism (Sachs et al., 2019; Kahn et al., 2022). In obesity, IMAT expansion correlates with skeletal muscle infiltration by T cells and macrophages and with impaired insulin sensitivity (Khan et al., 2015). Moreover, IMAT from obese individuals exhibits increased expression of proinflammatory cytokines, and factors secreted from IMAT have been shown to impair muscle insulin sensitivity, potentially through the release of inflammatory mediators, extracellular matrix (ECM) components, and increased local free fatty acid concentrations (Sachs et al., 2019). These findings indicate that IMAT expansion contributes to skeletal muscle insulin resistance and systemic metabolic dysfunction. IMAT displays transcriptomic profiles distinct from subcutaneous adipose tissue and skeletal muscle, with enrichment of pathways related to ECM remodeling, immune signaling, and stromal activation (Lutter et al., 2023; Chen et al., 2024). Emerging associations between IMAT and circulating proteomic and metabolic signatures further suggest that increased IMAT is linked to lower cognitive performance, particularly slower processing speed (Tanaka et al., 2025; Xu et al., 2025). Together, these observations position IMAT as a metabolically active and transcriptionally distinct adipose niche capable of exerting paracrine effects on skeletal muscle and potential endocrine influences.

The cellular origins of intermuscular adipose tissue (IMAT) are increasingly recognized as heterogeneous, involving fibro-adipogenic progenitors (FAPs) and other muscle-resident stem cell populations. FAPs are transcriptionally diverse stromal cells that support skeletal muscle homeostasis and regeneration but can expand and undergo dysregulated adipogenic differentiation under chronic inflammation or pathological conditions, thereby contributing to IMAT accumulation (Flores-Opazo et al., 2024; Uezumi et al., 2010). During normal muscle repair, FAP activation is transient and promotes regeneration through ECM remodeling and trophic interactions with muscle stem cells. However, persistent FAP activation in aging, metabolic disease, or muscle injury drives adipogenic differentiation and fibrotic remodeling within the skeletal muscle niche (Buras et al, 2019; Biferali et al., 2019). Fat cells surrounding the muscle bundles could also derive from mesenchymal progenitors such as mesenchymal stem cells or muscle-derived stem cells (SCs), which under altered differentiation cues adopt adipocyte-like features expressing PPARγ (De Coppi et al., 2006). This muscle-to-fat transition, driven by metabolic and environmental stressors such as obesity, hyperglycemia, hypoxia, and aging, engages PPAR, WNT, myokine, mitochondrial ROS, and PKCβ signaling (Vettor et al., 2009; Wang et al., 2024). In conclusion, IMAT expansion arises from aberrant lineage switching among FAPs and mesenchymal stem cells and targeting the pathways governing this fate transition may help preserve muscle integrity and mitigate sarcopenia and metabolic disease. Although IMAT expansion is strongly linked to metabolic disease, detailed cellular composition, transcriptional regulation, and interactions of human IMAT with skeletal muscle remain largely unexplored.

Building on our previous work demonstrating that human IMAT displays a gene expression profile distinct from skeletal muscle and subcutaneous fat (Sachs et al., 2019) and identifying IMAT-selective predictors of metabolic disease (Lutter et al., 2023), the present study aims to define drivers of IMAT expansion by focusing on a discrete gene expression signature selectively upregulated in IMAT. Using spatial transcriptomics combined with protein immunofluorescence in a rodent model of diet-induced cardiometabolic disease, we localized this signature within fibroadipogenic and stromal compartments and identified a transcription factor-driven pathway involving adipogenic transcription factor early B-cell factor 2 (EBF2) activation that mediates the conversion of human myoblasts into adipocyte-like cells. By resolving the spatial and transcriptional landscape of IMAT, our study provides insights into pathways that define IMAT identity and plasticity, offering a framework to understand how this depot contributes to muscle remodeling and the metabolic complications of cardiometabolic disease.

## Results

### Human IMAT-enriched gene expression signature spans brown/beige adipocyte biology, adipose niche remodeling, vascular/immune, and developmental functions

To identify potential drivers of human IMAT biology, we analyzed the genes selectively enriched in bulk RNA-seq datasets of IMAT compared with subcutaneous white adipose tissue and skeletal muscle from lean individuals. These genes were visualized in a heatmap of human IMAT-specific transcripts differentially expressed between IMAT and muscle or subcutaneous fat (Fig. 1A; Fig. S1A; ANOVA p_FDR < 0.01). Genes comprising the human IMAT-enriched signature were grouped, based on a manually curated literature search into several functional categories reflecting the complex stromal-vascular microenvironment of this depot. First, the presence of brown/beige adipocyte lineage and developmental regulators, including EBF2, PTGIS, SHOX2, TBX2, LRRC17, and RERGL, suggests activation of adipocyte lineage specification pathways (Fig 1A; genes highlighted in brown). EBF2, a transcription factor that promotes myoblast-to-brown adipocyte conversion and brown/beige lineage programs (Wang et al., 2014; Rajakumari et al., 2013) and PTGIS, which enhances adipogenesis and browning via PGI₂-IP receptor signaling (Massiera et al, 2003), highlight pathways associated with adipocyte recruitment. SHOX2, a homeobox transcription factor involved in mesenchymal patterning and depot-specific adipose biology (Lee et al., 2013), and the T-box transcription factor TBX2, which regulates developmental lineage specification and mesenchymal cell fate programs (Naiche et al., 2005), further support the presence of developmental regulatory pathways within human IMAT. LRRC17, a secreted leucine-rich repeat protein involved in stromal cell signaling and differentiation (Kim et al., 2009), and RERGL, a Ras-related GTP-binding protein implicated in growth regulation and cellular differentiation (Finlin et al., 2001), may likewise reflect stromal progenitor or lineage-specifying programs active within the IMAT niche.

**Figure 1.**
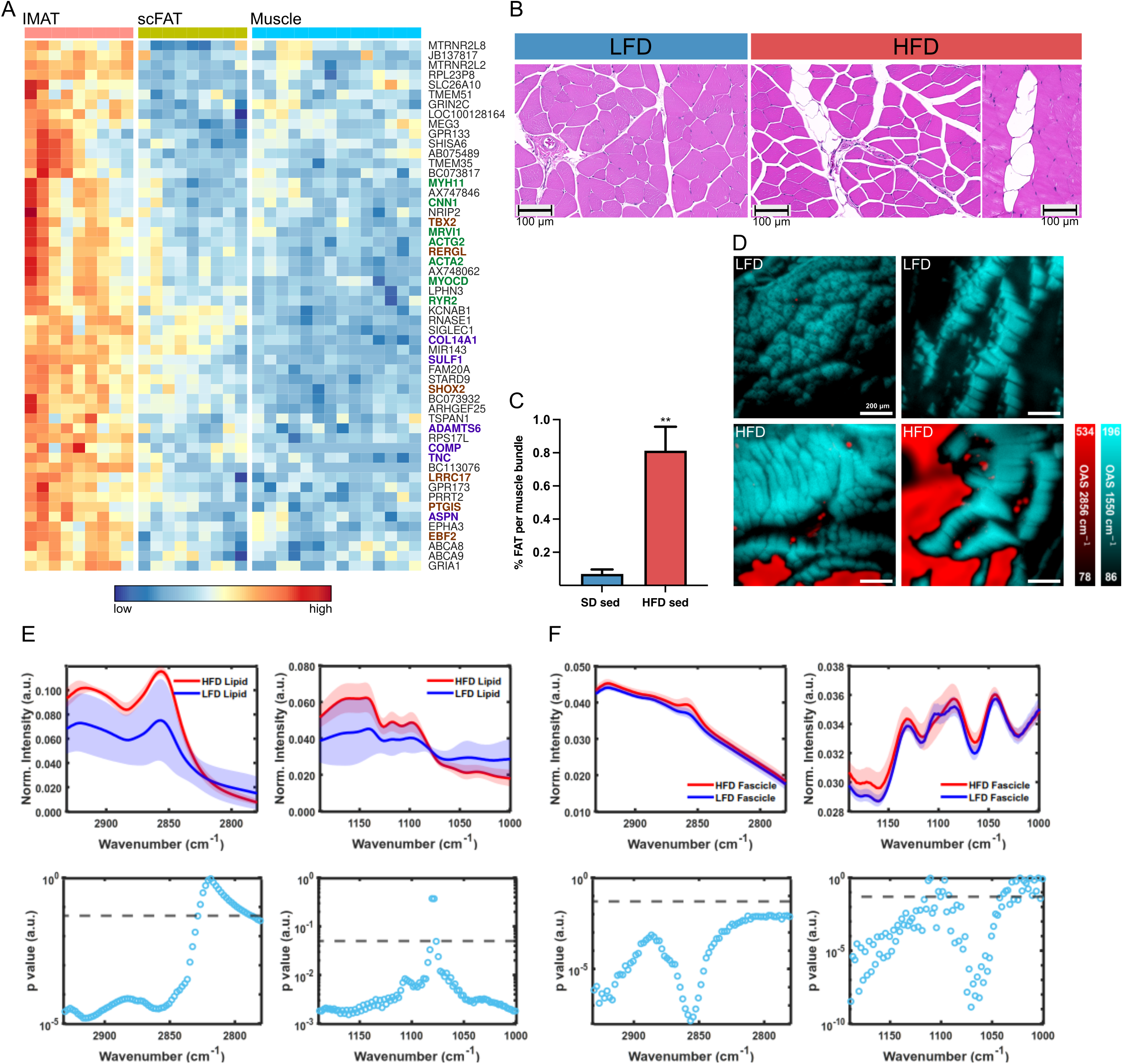
A) Heatmap of human IMAT specific genes differentially expressed between IMAT and muscle/subcutaneous FAT tissue of lean individuals (AVOVA p_fdr < 0.01) B) Representative histological sections of muscle quadriceps in high-fat-diet (HFD) and low-fat-diet (LFD) male mice. Intermuscular adipocytes are indicated by arrows. Scale bar 100um. C) Quantitative digital imaging analysis of histological sections in male mice (n = 7-8) after 84 days of HFD and LFD feeding. Data are presented as mean ± SEM. **P <0,0001 D) OA micrographs (1 mm × 1 mm) obtained from two channels (at 1550, 2856 cm^-1^) by mid-infrared optoacoustic microscopy (MiROM) (optoacoustic signal – OAS). E to F) OA spectra of lipid droplets (E) and muscle fibers (F) from HFD and LFD quadriceps muscle tissue (randomly selected spectral points; 20 points at lipid droplets and 40 points at muscle fibers) and p-values of t-tests on OA spectra for each wavenumber. The dashed line indicates p=0.05 which is considered as a statistically significant difference threshold. p < 0.05 is considered as a statistically significant difference.

A second group comprised smooth muscle and perivascular contractile genes (ACTA2, ACTG2, CNN1, MYH11, MYOCD, MRVI1, RYR2) here shown in green (Fig. 1A), suggesting contributions from vascular smooth muscle cells or mural cell populations. Consistent with this interpretation, lineage-tracing studies demonstrate that MYH11⁺ mural cells can give rise to adipocytes, particularly beige adipocytes during adipose remodeling (Long et al., 2014). ACTA2 represents a canonical contractile marker that is also induced in fibrotic adipose tissue (Eisinger et al., 2024). MYOCD, a coactivator of serum response factor, regulates smooth muscle lineage programs and mural cell plasticity (Pipes et al., 2006). The additional genes ACTG2, CNN1, MRVI1, and RYR2, which encode components of smooth-muscle contractile (Owens et al., 2004) and calcium-signaling machinery (Berridge, 2008), are consistent with a contractile mural cell transcriptional program but are not commonly used as adipocyte progenitor lineage markers and lack direct links to adipose tissue biology (Berry et al., 2013).

A third functional group included ECM and stromal remodeling genes ASPN, COL14A1, COMP, TNC, ADAMTS6, SULF1 (Fig. 1A, highlighted in purple), indicating active matrix organization and tissue remodeling within IMAT. ASPN modulates TGF-β signaling (Huang et al., 2022) and has recently been identified as an adipogenic gene in single-cell transcriptomic analyses (Tao et al., 2025). COL14A1, COMP, TNC, ADAMTS6, and SULF1 are components of ECM organization and regulators of growth factor signaling within the adipose microenvironment (Chiquet-Ehrismann et al., 2003; Mead et al., 2018; Ai et al., 2003; Theocharis et al., 2016).

Additional functional groups reflected the broader cellular complexity of the IMAT niche, including neuronal and synaptic signaling genes such as GRIA1, GRIN2C, LPHN3, PRRT2, SHISA6, KCNAB1, GPR173, GPR133 involved in glutamatergic neurotransmission and GPCR signaling within neuro-adipose circuits (Bartness et al., 2010; Zeng et al., 2015). Immune-associated genes of this IMAT signature, including the macrophage marker SIGLEC1 (Crocker et al., 2007) and the endothelial ribonuclease RNASE1 (Fischer et al., 2011), indicate immune and vascular proportions within IMAT. Enrichment of lipid transport genes ABCA8 and ABCA9 suggest activation of cholesterol and lipid trafficking pathways consistent with metabolic remodeling within the IMAT microenvironment (Tarling et al., 2013). A further set of signaling and membrane-associated genes, including EPHA3 (Pasquale, 2005) and TSPAN1 (Dharan et al., 2024), indicates membrane remodeling and tissue reorganization within the IMAT stromal niche, whereas TMEM35, TMEM51, STARD9, and NRIP2 remain insufficiently characterized in adipose biology. Among the noncoding transcripts, MIR143 (Esau et al., 2004) regulates adipocyte differentiation and MEG3 (Li et al., 2017) influences adipogenic versus osteogenic differentiation of adipose-derived mesenchymal stem cells. MTRNR2L2 and MTRNR2L8 represent humanin-like mitochondrial peptides associated with cytoprotective stress responses (Bodzioch et al., 2009).

Finally, several poorly annotated transcripts (AB075489, AX747846, AX748062, BC073817, BC073932, BC113076, JB137817, LOC100128164, RPL23P8, RPS17L) were enriched in IMAT, suggesting broader activation of noncoding and regulatory transcriptional programs potentially accompanying structural remodeling and cellular heterogeneity within the IMAT niche.

To further define the signaling networks associated with the human IMAT-enriched gene signature, we performed Ingenuity Pathway Analysis (IPA) (Supplementary Table 1). Among the 57 significantly enriched pathways we identified 10 canonical pathways related to adipocyte biology such as ECM organization, ILK and integrin signaling pathways (Fig. S1B and S1C; SupplementaryTable 1; Sun et al., 2013; Legate et al., 2009). Actin cytoskeleton signaling together with RHOA and Rho GTPase-ROCK pathways were also enriched, indicating activation of mechanotransduction processes linked to adipogenic lineage commitment and differentiation (McBeath et al., 2004; Wei et al., 2022). In addition, metabolic pathways including regulation of insulin-like growth factor (IGF) transport by IGF-binding proteins (Clemmons et al., 2012) and prostanoid biosynthesis (Sasaki et al., 2021) were enriched, consistent with signaling mechanisms that regulate adipocyte proliferation, differentiation, and lipid metabolism during IMAT expansion.

Together, gene- and pathway-level analyses define IMAT as a transcriptionally active niche integrating adipocyte lineage regulation, vascular mural progenitors, extracellular matrix remodeling, and metabolic signaling networks, alongside neural, immune, and regulatory components. These findings establish human IMAT as an actively remodeling stromal compartment that drives adipocyte recruitment and differentiation within skeletal muscle, rather than a passive reservoir of mature adipocytes.

### Diet-induced obesity promotes expansion of IMAT in male mice

To spatially define hotspots of IMAT expansion within skeletal muscle and explore specific changes in IMAT-associated genes related to cardiometabolic disease, we investigated skeletal muscles from diet-induced obese C57BL/6J mice. After 20 weeks of chronic 60% kcal high fat diet (HFD) these mice were hyperglycemic and glucose intolerant compared to control littermates fed a 10% kcal low fat diet (LFD) (ToKarz et al., 2021). Histological analysis of quadriceps revealed that HFD-induced IMAT expansion is predominantly visible around the muscle bundle (fascicles) at the perimysium level in male mice (Fig 1B and 1C) with female mice only showing marginal increases (Fig. S1D and S1E).

Using label-free mid-infrared optoacoustic microscopy (MiROM), which enables intrinsic biomolecular (e.g., lipids, proteins) characterization of biological tissue with minimal sample preparation (Pleitez et al., 2020; Ko et al., 2023; Kim, et al., 2026; Uluc et al., 2024, Gasparin et al., 2026a; Berger et al., 2025; Gasparin et al., 2026b) we confirmed and quantitatively assessed the expansion of IMAT in the quadriceps of HFD-fed and LFD-fed male mice (Fig. 1D-F). The mid-infrared (IR) absorption contrast micrograph obtained by MiROM, at 1150 cm-1 (Amide II band; N–H bending/C–N stretching), visualized protein distribution that represents muscle fascicles (Fig. 1F), and the micrograph at 2856 cm-1 (CH_2_ stretching) revealed the presence of lipid droplets (Fig. 1E). Accumulation of lipid droplets between muscle fascicles is mainly observed in HFD-fed male mice. Moreover, localized spectral analysis of lipid droplets and muscle fascicles has shown distinct spectroscopic features between the two groups. The upper panels of Fig. 1E showed averaged mid-IR spectra obtained from 20 lipid droplets in the quadriceps of HFD- and LFD-fed male mice, respectively. Within the CH_2_ vibrational region (2932-2780 cm^-1^) and the CO vibrational region (1190-1000 cm^-1^), which are relevant for lipid characterization, the area under the curve (AUC) of normalized spectral intensity was c.a. 1.4-fold and 1.1-fold higher in HFD-fed mice. The results indicated higher lipid content in HFD-fed mice compared to LFD-fed mice. Consistently, the lower panels of Fig. 1E showed that the spectral intensity differences between groups are statistically significant across the wavenumbers. Not only lipid droplets but also muscle fascicles exhibited distinct spectral features between the two groups. The upper panels of Fig. 1F show averaged mid-IR spectra obtained from 40 muscle fascicles in the quadriceps of HFD- and LFD-fed male mice, respectively. Although the differences are less significant than those observed for lipid droplets, muscle fascicles from HFD-fed mice show slightly higher intensity at the CH_2_ vibrational region (2932-2780 cm^-1^) and the CO vibrational region (1190-1000 cm^-1^) and also showed statistically significant differences in spectral intensity (the lower panels of Fig. 1F). Additionally, a peak at 1104 cm^-1^, relevant for lipid or carbohydrate characterization, was detected only in LFD-fed mice. These findings indicate that the biochemical composition of muscle fascicles also differs between HFD- and LFD-fed groups. Overall, through label-free assessment using MiROM, biochemical compositional differences in lipid droplets and muscle fascicles were observed between the two groups.

Integration of spatial transcriptomics and imaging reveals discrete IMAT niches and stromal enrichment of IMAT gene expression during diet-induced expansion. To map IMAT expansion within the muscle architecture, we integrated spatial transcriptomics with immunofluorescence staining for perilipin (adipocytes) and laminin (muscle fiber boundaries) on matched quadriceps sections from HFD- and LFD-fed male mice. This approach enabled spatially resolved analysis of molecular pathways and intercellular crosstalk between muscle and adipose compartments. Immunofluorescence staining of quadriceps muscle revealed an increase in the adipocyte specific perilipin signal in HFD-fed male mice compared with LFD-fed controls (Fig 2A). Quantitative digital imaging analysis of perilipin immunofluorescence demonstrated a significant increase in perilipin-positive cell niches surrounding muscle fascicles in HFD-fed mice, indicating lipid accumulation and expansion of IMAT in response to HFD (Fig 2B). Spatial cell-type composition was inferred using established cell-type deconvolution methods (Elosua-Bayes et al., 2021) in conjunction with a publicly available single-nucleus RNA-sequencing (snRNA-seq) reference dataset from mouse skeletal muscle (Ham et al., 2025). Based on the cell-type reference from snRNA-seq dataset (Fig. S2A) and marker genes (Fig. S2B), we inferred relative cell-type composition of each spatial transcriptomics spot (Fig. S2C). Although adipocytes and FAPs were among the cell types with the lowest overall proportions in both diets, both cell types significantly increased their proportion under HFD (Fig S2D). Regarding their spatial distributions we found adipocytes forming localized spatial clusters (Fig. 2C), whereas FAP-associated gene expression was more diffusely distributed across the tissue and was not modulated by HFD (Fig. 2D).

**Figure 2.**
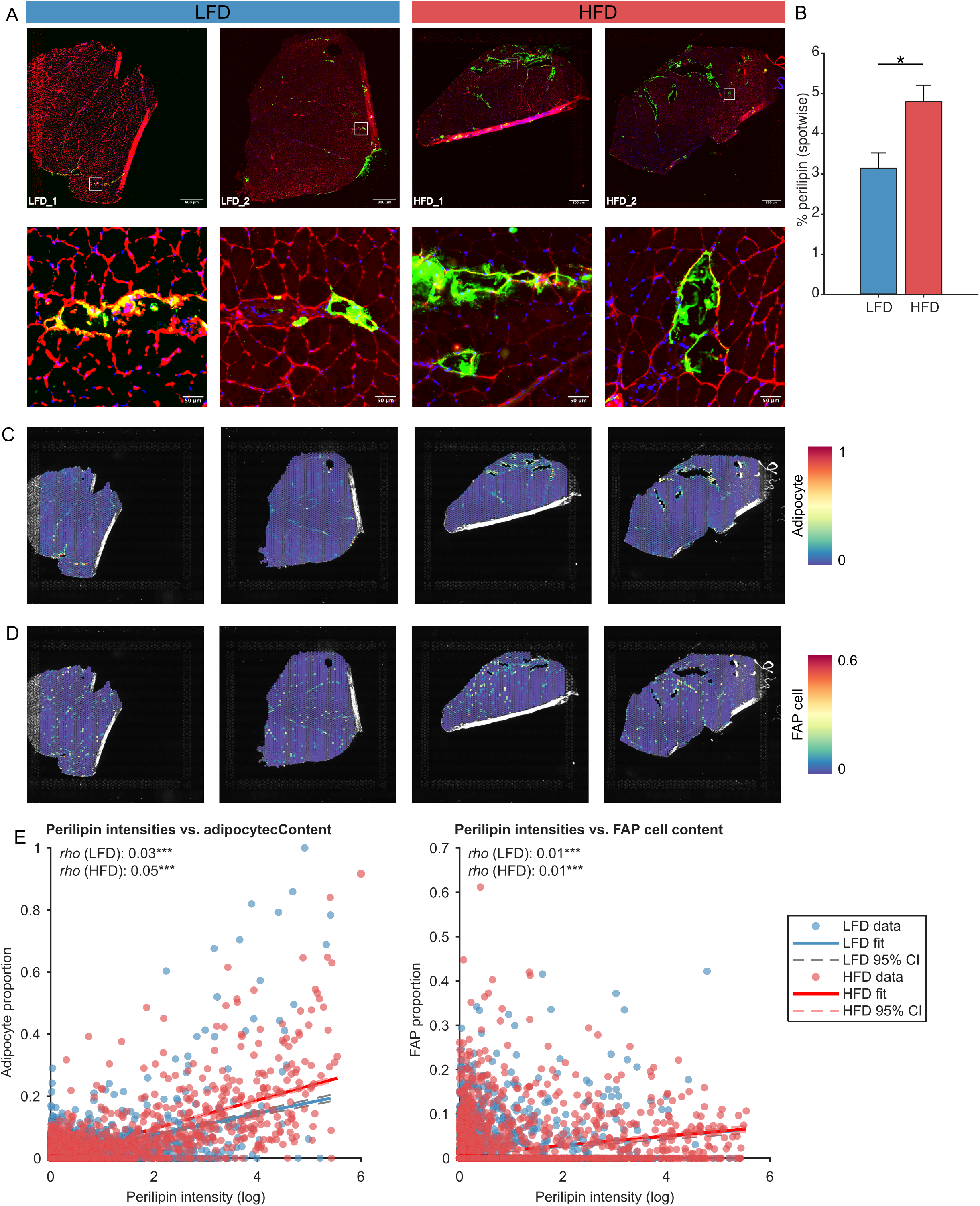
A) Immunofluorescence staining of laminin (red), perilipin (green), and DAPI (blue) in murine skeletal muscle cross-sections from low-fat diet (LFD) and high-fat diet (HFD) fed male C57BL/6 mice, acquired on Visium 10x Genomics slides (n = 2). B) Barplot mean and standard deviation of the spotwise perilipin staining percentage. Data are presented as mean ± SEM. *P <0,01 C) Relative spotwise adipocyte content estimated though deconvolution in LFD and HFD fed male mice of panel A. D) Relative spotwise FAP content estimated though deconvolution in LFD and HFD fed male mice of panel A. E) Scatterplot comparing spotwise perilipin intensity (mean) with adipocyte (left) and FAP (right) content for LFD (blue) and HFD (red) fed male mice of panel A. Correlation lines with 95% confidence intervals are shown for both diets.

To validate IMAT spot identification based on transcriptomic signatures, we performed spot-wise integration of immunofluorescence and spatial transcriptomics data. Perilipin fluorescence intensity strongly correlated with adipocyte-specific gene expression (Fig. 2E), confirming that transcriptional profiles accurately identify IMAT-containing regions. Consistent with this, HFD feeding significantly increased both perilipin staining intensity and adipocyte gene expression at the spot level, indicating enhanced adipocyte accumulation. FAP-associated gene expression did not correlate with perilipin intensity (Fig.E), highlighting a spatial and molecular dissociation from lipid-rich IMAT regions. Perilipin intensity was also increased in spots containing macrophages and smooth muscle cells (Fig. S2E), both of which are known to accumulate lipids, either as lipid-laden macrophages (Son et al., 2016) or through transdifferentiation of smooth muscle cell progenitors into beige adipocytes (Long et al., 2014).

### Unsupervised spatial clustering identifies adipocyte- and FAP-enriched niches with distinct remodeling states during high fat diet induced IMAT expansion

Unsupervised Louvain clustering of transcription spots identified nine distinct clusters (Fig. 3A) with varying abundance in relation to specific cell type distributions (Fig. S3A). Among this we identified Cluster 6 as the one with the highest adipocyte content (Fig 3B and S3A) and the Clusters 4, 6, 7 and 8 with the highest FAP content. (Fig 3C and S3A). Within these clusters we found a consistent increase in adipocyte (Cluster 6) or FAP content (4,6,7) under HFD except for cluster 8 where FAP content decreased. Cluster 8 was also characterized as the cluster with the highest pericyte content but in total with a minimum of spot number under both diets (Fig S3A and S3B). Over all clusters cell type reference gene markers also confirmed highest adipocyte content in cluster 6 and FAP content in the clusters 4,6,7, and 8 (Fig S3C and S3D). Macrophages were detected at low proportions across most clusters but were more prominent in clusters 4 and 6 (Fig S3A). In cluster 6, macrophage-associated gene expression further increased upon HFD feeding (Fig. S3A). Following these observations, we continued investigating the three clusters 4,6 and 7 for their spatial reorganization upon low and high fat diet.

**Figure 3.**
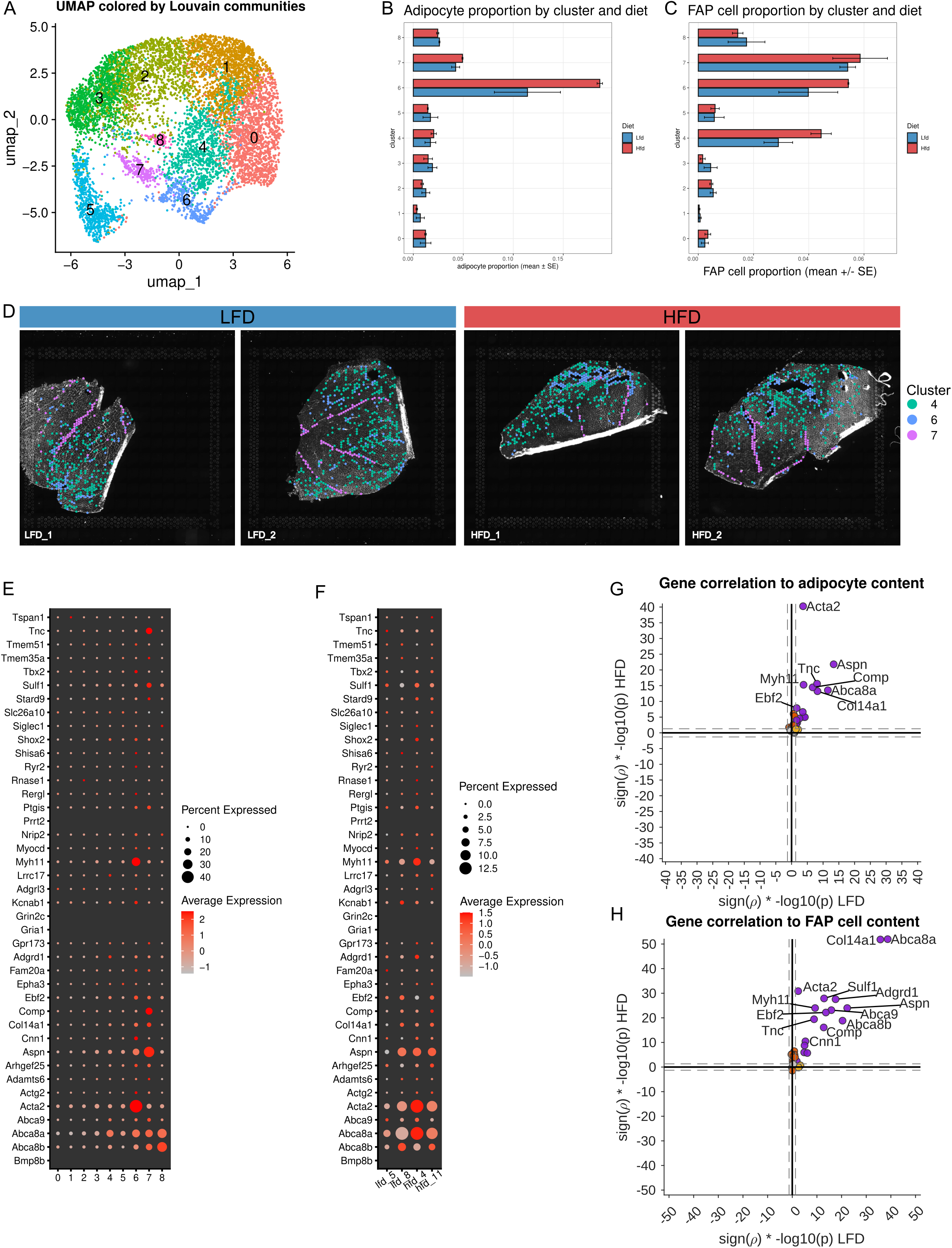
A) UMAP plot of all integrated spots with louvain clustering from low-fat diet (LFD) and high-fat diet (HFD) fed male C57BL/6 mice, acquired on Visium 10x Genomics slides (n = 2). Identifierd clusters are numbered from 0 to 8 and color coded. B) Distribution of adipocyte type proportion over all 9 identified spot clusters from LFD and HFD fed male mice. C) Distribution of FAP type proportion over all 9 identified spot clusters from LFD and HFD fed male mice. D) Spatial distribution of spots belonging to the three selected clusters 4, 6, and 7 for LFD and HFD fed male mice, shown on Visium 10x Genomics slides (n = 2). E) Dot plot for expression levels of the human IMAT specific gene signature over all 9 clusters determined in LFD and HFD fed male mice. F) Expression of the human IMAT specific gene signature between LFD and HFD fed male mice (n = 2). G-H) Scatterplot of directed p-values (p * sign (rho)) comparing IMAT specific correlation between spot wise human IMAT gene expression and cell type proportion for adipocytes (G) and FAPs (H) in LFD fed (x-axis) and HFD (y-axis) fed male mice. Purple dots denote genes significantly correlated in both directions; orange dots denote genes correlated in x-dimension (LFD) only and red in y-dimension (HFD) only.

Spatial spotwise distribution of cluster 4, 6, and 7 expression patterns revealed distinct spatial relationships (Fig. 3D). Cluster 6-associated gene expression was detected not only in spots with high adipocyte and FAP marker expression and strong perilipin staining, but also in adjacent, spatially proximal spots lacking clear adipocyte/FAP signatures or perilipin signal (Fig 2A and 2C). This distribution suggests that cluster 6 encompasses a heterogeneous cell population that includes, in addition to mature adipocytes, neighboring cell types such as FAPs, satellite cells, myogenic progenitors, or smooth muscle–like cells that may contribute to IMAT expansion. Consistent with this, Ingenuity Pathway Analysis (IPA) revealed upregulation of pathways related to adipocyte development, metabolism, and bioenergetics, as well as ECM remodeling, and inflammation (Fig. S3E; Supplementary Table 2 & 3). Cluster 7 expression preferentially overlapped with spots exhibiting low adipocyte/FAP gene expression and minimal or absent perilipin signal, consistent with a population representing an early or non-committed adipogenic state (Fig 3D, 2A, 2D). These cells may correspond to progenitor populations with the potential to differentiate into IMAT under permissive conditions. IPA did not indicate HFD-dependent modulation of signaling pathways in cluster 7 (Fig. S3E). Cluster 4-positive spots were more abundant and broadly distributed compared to clusters 6 and 7 (Fig. 3D), likely reflecting a more heterogeneous cellular composition (Fig. S3A). Within FAP-containing spots, cluster 4-associated gene expression was increased under HFD conditions (Fig 3C), suggesting a shift in cellular state or function in response to dietary challenge. IPA of cluster 4 genes indicated enrichment of inflammatory signaling pathways (Fig. S3E), consistent with the observed overlap between cluster 4 and macrophage-associated gene signatures (Fig. S3A).

### Partial cross-species conservation of the IMAT transcriptional program revealed by murine spatial and human bulk transcriptomics

To assess conservation of the human IMAT gene expression signature in mice, we mapped the human IMAT signature genes to mouse orthologs. Of these, 41 were successfully mapped and detected in the murine spatial transcriptomics dataset. The remaining 13 genes were excluded due to absent murine orthologs or insufficient functional annotation. The 41 conserved genes were subsequently mapped to the spatially resolved clusters identified in our dataset and we identified some to be slightly higher expressed in clusters 6 and 7 (Tnc, Sulf1, Myh11, Ebf2, Comp, Col14a1, Aspn, Acta2) (Fig. 3E) whereas only a slight increase in HFD could be detected for some genes (Acta2, Myh11, Abca8b; Fig. 3F). Consistently, correlating spotwise gene expression of the 41 IMAT genes with cell type composition across both diets indicated a significant increase in adipocyte, FAP, endothelial and smooth muscle cell proportions under HFD and at the same time we detected a decrease in myofibers and satellite cell proportions (Fig. S4A, S4B). In more detail, the majority of genes showed significant correlations with adipocytes (Fig. 3G) and FAPs (Fig. 3H) under both dietary conditions.

To extend these findings to a clinical setting, we analyzed human bulk RNA-seq data of IMAT biopsies from lean, obese, and T2D patients published previously (Sachs et al., 2019). All IMAT signature genes were detected, albeit with heterogeneous expression levels. Several genes (e.g., SHOX2, TSPAN1, ACAT2, CNN1, EBF2, ADAMTS6, SIGLEC1) showed a trend toward increased expression in obese and T2D patients compared to lean participants, although these differences did not reach statistical significance (Fig. S4C). Together with our murine spatial transcriptomics data, these results support partial conservation of the IMAT transcriptional signature across species. The comparatively modest and variable signal observed in bulk RNA-seq likely reflects limited sensitivity to detect spatially restricted and cell type-specific transcriptional changes, underscoring the advantage of spatial transcriptomics for resolving IMAT expansion in situ.

### EBF2 promotes trans-differentiation of human myoblasts toward an adipogenic phenotype and may contribute to IMAT expansion in diet-induced obesity

Because EBF2 was consistently enriched in human and murine IMAT in our RNA-seq analyses (Fig. 1A, 3E–H, S4C) and has been linked to myoblast-to-brown adipocyte conversion in mice (Wang et al., 2011), we investigated whether it contributes to IMAT expansion in cardiometabolic disease. In an independent human cohort (see methods), EBF2 was markedly upregulated in IMAT versus skeletal muscle (Fig. 4A), validating our bulk RNA-seq findings (Fig. 1A). PPARδ and MYF5 were included in this analysis as markers of adipogenic and myogenic lineage commitment, respectively, to contextualize EBF2 expression within relevant differentiation pathways. In addition, we observed slight, although not significant increases in EBF2 expression levels in type 2 diabetes subjects compared to healthy control participants (Fig. S4C).

**Figure 4.**
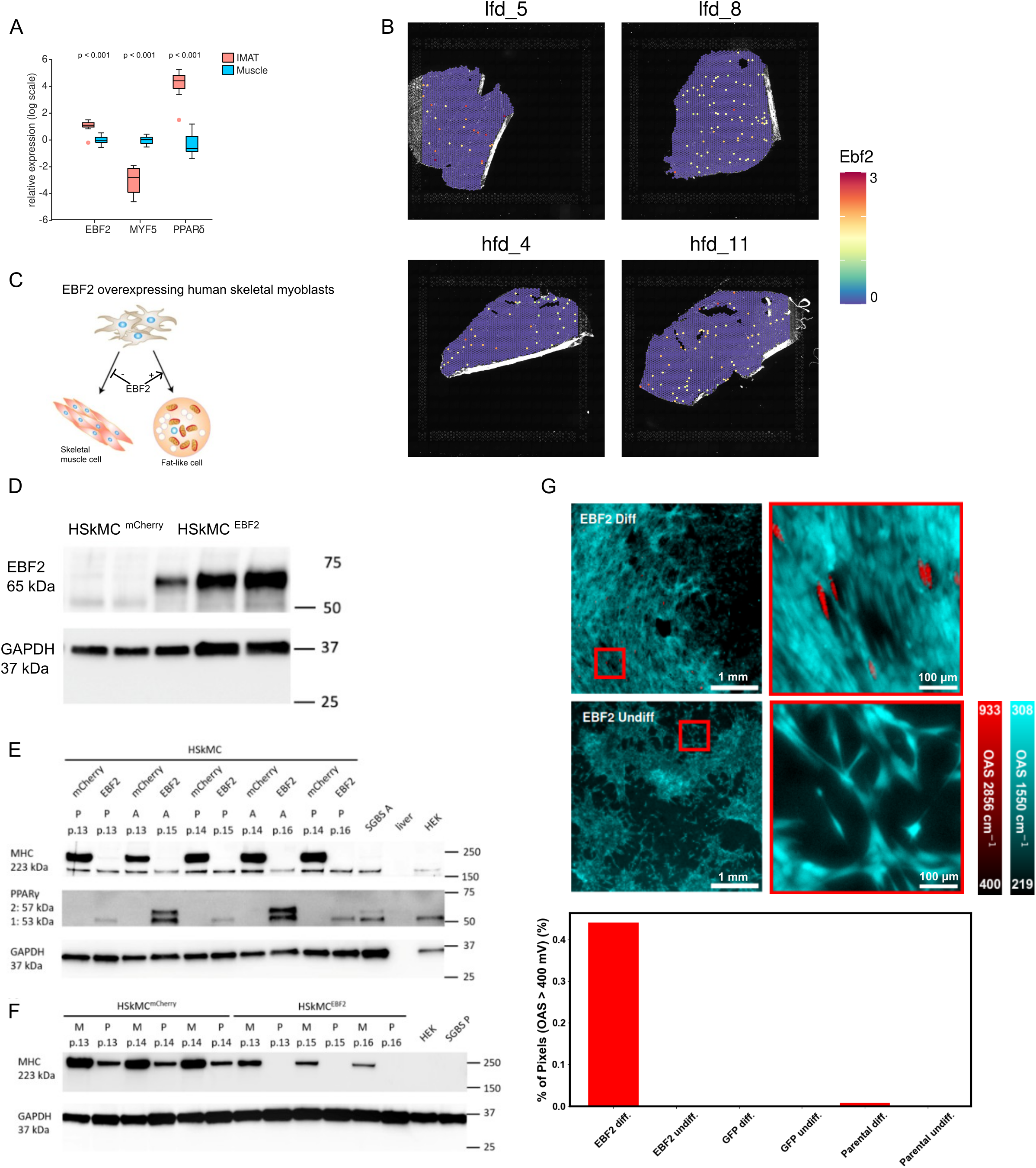
A) Expression levels of EBF2, PPARδ, and MYF5 were quantified by RT-PCR in human intermuscular adipose tissue (IMAT) and skeletal muscle (n = 22/30 (IMAT/Muscle). Data are presented as mean ± SEM; statistical significance was determined by unpaired t-test. B) Spatial expression of EBF2 in low-fat diet (LFD) and high-fat diet (HFD) fed male C57BL/6 mice acquired on Visium 10x Genomics slides (n = 2). C) Schematic overview of the experimental design. Human primary myoblasts were transduced with lentiviral vectors encoding EBF2 (HSkMCEBF2) or a control mCherry-GFP (HSkMCmCherry) construct to test whether EBF2 overexpression induces transdifferentiation toward a fat-like phenotype and affects myogenic or adipogenic differentiation. D) Western blotting of EBF2 in HSkMCEBF2 and HSkMCmCherry cells after lentiviral transduction and transdifferentiation with adipogenic media. GAPDH was used as loading control. 5 µg proteins were loaded per sample. Size (in kDa) of molecular weight marker is indicated on the right. E) Comparison between adipocenic transdifferentiated and proliferating HSkMCmCherry and HSkMCEBF2 cells. Western blotting of MHC in HSkMCEBF2 and HSkMCmCherry after 14 days of adipogenic differentiation (A) or proliferation (P). GAPDH was used as loading control. 5 µg proteins were loaded per sample. Size (in kDa) of molecular weight marker is indicated on the right., HEK = HEK 293T cells, Liver homogenate, SGBS = SGBS cells, P = proliferating, A = adipogenic differentiated, p. = passage number. F) Comparison between myogenic differentiated and proliferating HSkMCmCherry and HSkMCEBF2 cells. Western blotting of MHC in HSkMCEBF2 and HSkMCmCherry after 7 days of myogenic differentiation (M) or proliferation (P). GAPDH was used as loading control. 5 µg proteins were loaded per sample. Size (in kDa) of molecular weight marker is indicated on the right., HEK = HEK 293T cells, SGBS = SGBS cells, P = proliferating, M = myogenic differentiated, p. = passage number. G) Human skeletal muscle cell (HSkMC) adipogenic differentiation (Diff., (left panel) and non-differentiation (Non-Diff. right panel) comparative analysis by mid-infrared optoacoustic microscopy (MiROM). OA micrograph of lentiviral EBF2 gene overexpression in HSkMC, Big FOV (4 mm × 4 mm) is on the left side, and small FOV (500 μm×500 μm) is on the right side in two channels: red (at 2856 cm^-1^; CH_2_ band - symmetric CH_2_ stretching) and cyan (at 1550 cm^-1^; Amide II band – N-H bending/ C-N stretching). Box plot indicates the comparison between the percentage of pixels in which optoacoustic signal (OAS) is higher than 400 mV in a big FOV at 2856 cm^-1^, which provides the predominant contrast of lipid droplets.

Spatial distribution indicates that Ebf2 expression is a consistent feature of adipogenic and FAP cell niches, with a modest trend toward increased Ebf2 expression under HFD conditions (Fig. 4B) that may contribute to IMAT expansion. Analysis of genes colocalizing with Ebf2-expressing spots revealed coordinated HFD-dependent transcriptional changes (Fig S5A). HFD induced genes linked to lipid handling and adipogenic potential, including Adipor1, Lpar1, and Fabp4, alongside reduced expression of Cd36, indicating altered fatty acid uptake and intracellular lipid trafficking (Yamauchi et al., 2002; Febbraio et al., 1999; Hotamisligil et al., 1996). Within this framework, Ebf2 acts as an upstream organizer of adipogenic transcriptional programs, with PPARγ as the direct transcriptional driver and Fabp4 and Cd36 representing downstream effector genes. Concurrent reduced expression of oxidative metabolism genes (Ldhb, Phkb, Slc25a37) alongside increased Fbxo32, a marker of protein degradation and atrophy (Bodine et al., 2001), and decreased stress-response and ECM genes (Hspa5, Sparc) (Bradshaw et al., 2003) indicate impaired energy metabolism and disrupted cellular stress and tissue organization. Collectively, these changes define an Ebf2-associated program under HFD that promotes a lipid-enriched environment with impaired energy metabolism that favors IMAT expansion and dysfunction.

To further characterize IMAT-associated transcriptional networks, we analyzed the expression of STRING-identified Ebf2 interaction genes across all nine clusters under LFD and HFD conditions (Fig. S5B). Focusing on IMAT-enriched clusters (clusters 4, 6, and 7), we observed cluster- and diet-specific activation patterns. In cluster 4, Hnrnpu, a regulator of RNA processing and chromatin organization, was detected under both conditions, whereas HFD induced additional interaction partners, including Zfp423, a marker of adipogenic lineage commitment, as well as Ebf3 and Zfp516, which are associated with adipogenic and thermogenic transcriptional programs (Chen et al., 2022). In cluster 6, Ebf2-associated genes under LFD included Ehmt1, an epigenetic regulator of adipocyte differentiation (Ohno et al., 2013), together with Hnrnpu and Id1, a factor linked to progenitor states (Chen et al., 2022). Under HFD, cluster 6 exhibited induction of a broader adipogenic program, including Cidea and the key adipogenic regulators Pparγ and Prdm16, indicative of mature and metabolically active adipocytes (Rosen et al., 1999; Seale et al., 2011), alongside Zfp516 (Chen et al., 2022). In cluster 7, HFD promoted expression of Ehmt1, Hnrnpu, Id1, and Paqr9, suggesting a more transitional or metabolically responsive cellular state (Lin et al., 2021). In conclusion, HFD induced expression of multiple Ebf2-interacting genes, most prominently in cluster 6, where key regulators of adipocyte differentiation and metabolism were upregulated. This identifies cluster 6 as a central node of IMAT expansion driven by Ebf2-associated programs, while clusters 4 and 7 likely represent earlier or transitional states along this trajectory.

To directly assess the role of EBF2 in driving adipogenic reprogramming, human primary myoblasts were transduced with lentiviral vectors encoding EBF2 or a control mCherry-GFP construct (scheme in Fig 4C). Western blot analysis confirmed successful EBF2 overexpression (Fig 4D) and revealed induction of the adipogenic marker PPARγ, while the myogenic marker myosin heavy chain (MHC) was reduced under adipogenic differentiation conditions (Fig. 4E). Under myogenic differentiation conditions, MHC expression was preserved (Fig. 4F) but reduced, indicating that EBF2 selectively promotes adipogenic but not myogenic differentiation pathways.

Label-free biomolecular imaging of human myoblasts was performed using MiROM, a bond-specific label-free imaging technique that detects molecular contrast without exogenous labels (Fig. 4G). Three human primary skeletal muscle myoblast cell lines, EBF2-overexpressing, GFP-overexpressing, and wild type, were imaged under two culture conditions: adipogenic differentiation (growth medium supplemented with an adipogenic cocktail) and non-differentiation (standard growth medium). MiROM measurements were conducted in two vibrational channels: 2856 cm⁻¹ (CH₂ symmetric stretching) for visualization of lipid droplets, reflecting the triglyceride-rich content of intracellular lipid stores, and 1550 cm⁻¹ (Amide II band; N–H bending/C–N stretching) for mapping the overall protein distribution representing myoblast structures. The micrographs obtained with MiROM, were acquired at both large (4 mm × 4 mm) and small (500 μm × 500 μm) fields of view (FOV) to capture cellular and subcellular features. Lipid accumulation was quantified based on signal intensity at 2856 cm⁻¹, where lipid droplets exhibited strong optoacoustic responses (>400 mV). The proportion of pixels exceeding 400 mV at large FOV was calculated to estimate lipid-rich regions, enabling comparative assessment of fat accumulation across cell lines and conditions. Lipid droplet formation was observed exclusively in EBF2-overexpressing myoblasts (Fig. 4G), whereas neither GFP-overexpressing nor wild-type cells exhibited detectable lipid accumulation (Fig. S5C). Quantitative analysis confirmed that the percentage of high-intensity lipid-associated pixels (>400 mV at 2856 cm⁻¹) exceeded 0.4% in EBF2-overexpressing cells, while wild-type and control primary human myoblasts remained near background levels (∼0%) (Fig. 4G). Collectively, these findings demonstrate that EBF2 overexpression in human myoblasts induces a trans-differentiation toward a fat-like phenotype, characterized by lipid droplet accumulation and activation of adipogenic markers, exhibiting some of the molecular and structural features of IMAT.

## Discussion

This study reveals conserved core mechanisms driving IMAT remodeling in cardiometabolic disease by combining spatial transcriptomic mapping in a murine model with cross-species identification of conserved transcriptional programs, cellular associations with adipocytes and fibro-adipogenic progenitors, and shared pathways linking ECM remodeling, inflammation, and adipogenesis in human IMAT. Importantly, functional validation in human primary myogenic cells identifies EBF2 as a key upstream regulator capable of driving adipogenic reprogramming, providing direct evidence that these conserved programs are not only associative but mechanistically linked to muscle remodeling and metabolic dysfunction.

Using spatial transcriptomics in a diet-induced cardiometabolic disease mouse model, we localized the human IMAT enriched gene signature to fibroadipogenic and stromal niches surrounding muscle fascicles of male mice. HFD enhanced the co-expression of adipogenic and stromal gene networks, indicating coordinated activation of these cell types during IMAT expansion. This spatially resolved approach allowed identification of cellular interactions within intact muscle architecture and provided mechanistic insight into how metabolic stress reshapes muscle tissue composition through localized adipogenic remodeling. The lack of correlation between FAP-associated gene expression and perilipin staining suggests that FAP abundance does not directly reflect local adipocyte formation within IMAT-containing regions. Instead, FAPs likely represent a spatially dispersed stromal progenitor population with heterogeneous differentiation potential, consistent with their established capacity to adopt adipogenic or fibrogenic fates depending on local microenvironmental cues (Uezumi et al., 2010; Chen et al., 2022). The spatial separation of FAP-rich and perilipin-positive regions suggests that progenitor presence is functionally distinct from mature adipocyte accumulation and may reflect IMAT remodeling or an intermediate, non-differentiating adipogenic state that does not directly contribute to HFD-induced IMAT expansion. Consistent with this, snRNA-seq studies have identified heterogeneous preadipocyte populations within IMAT, indicating that adipogenic progenitors are not uniformly committed to mature adipocyte formation (Zemski Berry et al., 2025). Importantly, FAP-to-adipocyte differentiation is a dynamic and context-dependent process, in which progenitor-associated gene expression is downregulated upon lipid accumulation and induction of mature adipocyte markers such as perilipin, reflecting progressive lineage commitment (Chen et al., 2022). Spatial transcriptomics is therefore well suited to capture distinct stages of this transition; however, as a snapshot-based approach, it may detect progenitor and differentiated states in spatially and temporally distinct locations. This temporal and spatial uncoupling may explain the absence of spot-wise correlation despite a potential lineage relationship between FAPs and IMAT adipocytes.

The translational relevance of these findings was supported by analysis of our human cohorts (Sachs et al., 2019), in which the human IMAT gene signature was slightly upregulated in IMAT from individuals with obesity or type 2 diabetes, paralleling the patterns observed in HFD fed mice. These cross-species concordances highlight conserved molecular pathways linking IMAT remodeling with inflammation, ECM dynamics, and metabolic dysregulation. Although the IMAT transcriptional signature showed partial conservation across species, this was incomplete, likely reflecting both biological differences and the limited resolution of bulk RNA-seq in human samples. The inability of bulk approaches to resolve spatially restricted and cell type specific signals likely underestimates IMAT complexity, whereas spatial transcriptomics enables direct identification of niche-specific programs in situ, providing a more refined view of IMAT organization and expansion. The enrichment of poorly annotated transcripts in the human IMAT signature points to previously unrecognized regulatory mechanisms underlying IMAT remodeling beyond established adipogenic pathways. Among the conserved IMAT-enriched genes, EBF2 emerged as a potential upstream regulator of adipogenic reprogramming. Although EBF2 expression showed only a modest, nonsignificant increase in IMAT from patients with type 2 diabetes, even subtle activation of this transcription factor may influence lineage commitment and critical time windows in the muscle. Our functional experiments demonstrate that EBF2 overexpression in human myoblasts induces adipogenic transdifferentiation, characterized by lipid accumulation and activation of adipogenic markers, suggesting that EBF2 can reprogram myogenic precursors toward a fat-like phenotype. Notably, the use of MiROM provided a powerful, label-free method to detect and characterize these fat-like cells with high spatial and chemical specificity. MiROM leverages lipid- and protein-specific vibrational contrasts to enable quantitative, non-invasive visualization of lipid droplet formation and metabolic remodeling in living or fixed cells (Pleitez et al., 2020; Ko et al., 2023). This technology surpasses traditional staining approaches in sensitivity and resolution, allowing early detection of adipogenic reprogramming and offering a scalable tool to study IMAT expansion under metabolic or therapeutic modulation.

A limitation of this study relates to tissue sectioning, which is inherently challenging due to the distinct mechanical and compositional properties of skeletal muscle and adipose tissue. Adipose-rich regions are difficult to process, often leading to tissue instability, tearing, or loss during sectioning (https://www.leicabiosystems.com/en-de/knowledge-pathway/processing-fatty-specimens/?utm_source). Consistent with this, we observed occasional detachment of adipose tissue from muscle, resulting in voids that were excluded from analysis. Although this reduced the number of analyzable IMAT regions, focusing on intact sections enabled robust characterization of IMAT niche organization. Notably, we identified spatial niches within muscle displaying adipocyte-like transcriptional profiles despite lacking overt adipose morphology. This raises the possibility that IMAT expansion may partly involve local conversion or transdifferentiation of resident muscle-associated cells. Future studies combining improved tissue preservation with longitudinal spatial transcriptomics and lineage-tracing approaches will be required to directly test these mechanisms and define the cellular origins of IMAT expansion.

In summary, our data establish IMAT as a transcriptionally and spatially distinct adipose niche within skeletal muscle, defined by a hybrid molecular identity integrating beige adipocyte-associated, stromal, vascular, and immune programs. Unlike classical adipose depots, IMAT resides in close proximity to muscle fibers and probably engages in reciprocal signaling with myogenic and stromal cells. Pathway analysis reveals coordinated activation of adipogenic reprogramming, extracellular matrix remodeling, and inflammatory signaling, highlighting its role as a dynamic regulator of tissue adaptation and metabolic homeostasis. Notably, the ability of EBF2 to induce adipogenic transdifferentiation in human primary myoblasts, together with spatial mapping of IMAT expansion, identifies lineage plasticity as a central mechanism of IMAT formation. We therefore propose that IMAT expansion arises from spatially restricted lineage transitions among FAPs and myogenic precursor cells. Collectively, these findings define IMAT as a metabolically active niche whose dysregulation contributes to muscle remodeling and cardiometabolic disease. Targeting these fate decisions may offer new strategies to preserve muscle integrity.

## Methods

### Study population

Bulk RNA-seq datasets from human skeletal muscle, intermuscular adipose tissue (IMAT), and subcutaneous adipose tissue (SAT) were obtained from previously published studies (Sachs et al. 2019; Lutter et al. 2023). All participants provided written informed consent, and study protocols were approved by the Colorado Multiple Institution Review Board at the University of Colorado. In brief, skeletal muscle, IMAT, and SAT biopsies were collected from lean, obese, and type 2 diabetic individuals, as well as from lean endurance-trained athletes and sedentary controls. Detailed participant characteristics, inclusion criteria, and clinical parameters have been published previously.

### Bulk RNA Sequencing Analysis in human samples

Bulk RNA-seq datasets used in this study were generated and described previously in detail (Sachs et al., 2019; Lutter et al., 2023) and were reprocessed here for comparative transcriptomic analyses.

### Mice

For IMAT expansion analysis male and female C57BL/6J mice were obtained from Charles River Laboratories (Germany). At 12 weeks of age, animals of each sex were stratified based on body weight and randomly assigned to either a standard diet (SD; 1310; Altromin Spezialfutter GmbH, Germany) or a high-fat diet (HFD; D12331, Research Diets INC, USA). Mice were maintained at a 12h light/dark cycle with ad libitum access to food and water for 12 weeks. At the end of the study, animals were euthanized under carbon dioxide anesthesia, and organs were collected. From each mouse, one quadriceps muscle was fixed in paraformaldehyde for histopathological examination. For spatial transcriptomics analysis seven month old male C57BL/6J mice were fed a low fat diet (LFD; 10 kcal% fat, 70 kcal% carbohydrate, 20 kcal% protein; 3.82 kcal/g; D12450Ji Research diets) or a high fat diet (60 kcal% fat, 20 kcal% carbohydrate, 20 kcal% protein; 5.21 kcal/g; D12492i Research diets) for 20 weeks. At study termination, mice were euthanized and quadriceps muscles carefully isolated and flash frozen in liquid nitrogen. All procedures were approved by the local Animal Use and Care Committee and the local authorities of Upper Bavaria, Germany in accordance with European and German animal welfare regulations.

### Histology with imaging analysis

Dissected m. quadriceps samples were fixed in 4% (w/v) neutrally buffered formalin, sectioned to obtain two cross-sections and one butterfly-shaped longitudinal section, and embedded in paraffin with the cut surfaces facing downward. Standard FFPE sections were cut at 3 µm thickness for H&E staining and scanned using an Axio Scan.Z1 digital slide scanner (Zeiss, Jena, Germany) equipped with a 20× objective. Images were evaluated using image analysis software Definiens Developer XD 2 (Definiens AG, Munich, Germany). Histopathological evaluation shown in Fig 1D, 1E, S1D and S1E was performed by a blinded, certified pathologist on randomly selected regions. Due to well-known technical challenges in adipose-muscle sectioning, occasional adipose detachment created voids that were excluded from analysis.

### Spatial transcriptomics

Fresh-frozen quadriceps muscles from male mice were used for spatial transcriptomic analysis. The workflow followed the 10x Genomics Visium Spatial Gene Expression protocol (Visium Spatial Gene Expression Slide & Reagent Kit, 4 rxns, PN-1000187). Briefly, quadriceps muscles were cryosectioned at the Helmholtz Munich Core Facility Pathology and Tissue Analytics (CF-PTA) into 10 µm-thick cross sections and placed within the fiducial frame of a Visium spatial gene expression slide. During sectioning occasional detachment of adipose tissue from the muscle was observed during sectioning as a known technical constraint, generating tissue voids that were excluded from subsequent analyses. Sectioning was followed by fixation and staining with DAPI for nuclei and with the following antibodies: rat anti-Laminin (1:200, ab11576, Abcam) a basement membrane marker used to clearly delineate individual myofibers for segmentation, and rabbit anti-Perilipin (1:100, #57117SF, Cell Signaling) used as a marker for adipocytes. The following secondary antibodies were applied for detection: donkey anti-rabbit AlexaFluor555 (1:100, A31572, Invitrogen) and goat anti-rat AlexaFluor647 (1:100, A21247, Invitrogen). Whole section images were acquired using an AxioScan 1 digital slide scanner (Zeiss, Oberkochen, Germany) equipped with a 20x objective. Exposure times, laser intensities, and filter settings were kept constant across all samples to ensure comparability. Subsequently, tissue sections were permeabilized for 18 minutes following the Visium Spatial Tissue Optimization workflow, allowing cellular mRNA to hybridize to oligonucleotide primers located on the spatial gene expression capture spots. cDNA libraries were generated from the captured transcripts and sequenced at the Helmholtz Munich Core Facility Genomics (CF-GEN). Spatial barcodes associated with each capture spot were then used to map sequencing reads back to the corresponding quadriceps tissue sections, enabling spatially resolved gene expression profiling as shown in Fig 2A to C, S2C, 3D and 4B.

### Spatial transcriptomics data processing

To generate the count matrices, FASTQ files and histology images for each sample were processed with Space Ranger (v3.0.1). Reads were aligned to the Mus musculus reference genome assembly (GRCm39, Ensembl release 110) using spaceranger count. Downstream analysis was performed using Seurat v5.3.0 (Hao et al. 2024) in R v4.4. Data were normalized using SCTransform (Hafemeister et al. 2019), and dimensionality reduction was performed. Dataset integration was carried out using Harmony (Korsunsky et al., 2019). First 13 principal components (PCs) were used to identify neighbours and clustering was done using louvain algorithm at resolution 0.6. SCTransform-normalized data were used for integration and clustering. For all other downstream analyses and visualizations, log-normalized expression values generated with NormalizeData were used. The data was visualized using Uniform Manifold Approximation and Reduction (UMAP).

### Spot wise deconvolution

To infer the cell type composition of spots, we used a reference-based method, SPOTlight (Elosua-Bayes et al., 2021). SPOTlight applies a seeded non-negative matrix factorization (NMF) regression approach using cell-type marker genes and non-negative least squares (NNLS) optimization to estimate the contribution of different cell types within each spatial transcriptomic spot. We used a mouse single-nuclei RNA-seq dataset of skeletal muscle as a reference (Ham et al., 2025). The snRNA-seq reference dataset was normalized using logNormCounts() from the scater package (McCarthy et al., 2017). Highly variable genes (HVGs) were identified using scran’s modelGeneVar() (Lun et al., 2016) on a feature set excluding ribosomal and mitochondrial genes, followed by getTopHVGs() to select the top 3,000 HVGs. Cell-type specific marker genes were identified for each annotated identity using the scoreMarkers() function from the scran package. Genes with a mean area under the curve (AUC) greater than 0.8 were used as markers for each cell type. To ensure balanced representation across cell identities, up to 130 cells per cell type were randomly selected to generate a balanced reference subset used for downstream deconvolution analyses.

### Cell culture

Human skeletal muscle cells (HSkMC; Cell Applications, Inc.) overexpressing either EBF2 (HSkMCEBF2) or mCherry (HSkMCmCherry) as control were cultured under antibiotic selection with 125 µg/mL Geneticin™ (G418 sulfate; Gibco™) or 0.4 µg/mL Puromycin (Sigma-Aldrich), respectively, according to the manufacturer’s protocol. Cells were maintained in HSkMC Growth Medium (Cell Applications, Inc.) at 37°C and 5% CO₂, with medium changes every two days. When cultures reached 85–95% confluency, cells were detached using 0.05% Trypsin-EDTA (Gibco™), centrifuged at 220 × g for 5 min, and reseeded at 5,000 cells/cm². For myogenic differentiation, HSkMC were seeded at 32,000 cells/cm², and differentiation was induced by replacing the growth medium with HSkMC Differentiation Medium (Cell Applications, Inc.), refreshed every second day until multinucleated myotubes appeared (day 7). For adipogenic differentiation, the protocol described by Berti et al. for Simpson–Golabi–Behmel syndrome (SGBS) cells was followed (Berti et a., 2015). Cells were cultured in DMEM/F-12 (1:1) + GlutaMAX™ (Gibco™) supplemented with 5% FBS and 1% Penicillin–Streptomycin (Gibco™). Differentiation was performed in two phases: an induction phase (7 days) and a differentiation phase (11 days). Both media contained Vitamin B5 (17 µmol/L), Biotin (1 µmol/L), Apo-Transferrin (2 µg/mL), Insulin (1 µmol/L; Gibco™), and Dexamethasone (1 µmol/L). The induction medium additionally included IBMX (0.5 mmol/L), Triiodo-L-thyronine (T3; 2 nmol/L), and Indomethacin (50 µmol/L), which were omitted from the subsequent differentiation medium. After 18 days, mature adipocyte-like cells with visible lipid droplet accumulation were obtained. SGBS preadipocytes served as positive controls and were cultured and differentiated using the same adipogenic protocol. Lysed HEK293T cells were used as protein controls for Western blot analyses.

### Mid-infrared optoacoustic microscopy (MiROM)

MiROM is a vibrational spectroscopic imaging technique that detects the chemical composition of a sample by acquiring sound generated by mid-infrared light absorbed by the sample. To assess the unique vibrational frequencies of molecules, MiROM uses a combination of mid-infrared (mid-IR) excitation and ultrasound acoustic detection. Based on the optoacoustic effect, biological samples absorb mid-IR radiation and convert its energy into heat. This localized heating causes thermoelastic expansion, generating acoustic waves that are linearly related to the absorption coefficient. Molecules absorb mid-IR radiation at resonant frequencies that match vibrational modes of molecules, and each molecule exhibits a unique set of vibrational modes. By tuning the excitation wavelength to these bond-selective vibrational transitions and detecting the resulting optoacoustic signals, MiROM provides intrinsic label-free chemical contrast in biological samples.

MiROM (Pleitez et al., 2020; Ko et al., 2023; Kim et al., 2026; Uluc et al., 2024, Gasparin et al., 2026a; Berger et al., 2025; Gasparin et al., 2026b) used in this work, operates in transmission mode and uses a broadband quantum-cascade laser (QCL, MIRcat, Daylight solution) covering 2932-910 cm^-1^ (3.4-11 µm) with a spectral width of ≤1 cm^-1^. To avoid interference from CO_2_ and water vapor in the mid-IR spectrum, the mid-IR beam path from the QCL has been purged with a constant flow of N_2_ gas. The pulse duration is 20 ns, and the repetition rate is 100 kHz. The laser beam is focused with an x36 reflective objective (Newport) with a numerical aperture of 0.5. The sample is located on the mid-IR transparent window (ZnS, Crystal). To acquire an optoacoustic signal, we use an ultrasound transducer (Imasonic) with a central frequency of 20 MHz, which is placed above the sample. The optical diffraction-limited focus by the objective and the transducer’s focus are coaxially aligned. The generated optoacoustic wave from mid-IR radiation at the optical focus travels through the coupling medium (deionized water) and is detected by the ultrasound transducer. The optoacoustic signal is acquired by a data acquisition card (DAQ, Gage Applied) after amplification by a 63 dB amplifier (MITEQ) and filtering with a 50 MHz low-pass filter (Mini-circuits). The micrograph is obtained using an xy-motorized stage (Physik Instrumente) through raster scanning. Data has been collected by MATLAB 2018b.

### Statistical Analysis

Bulk RNA Seq processing and statistical analysis was performed as described previously (Sachs et al., 2019; Lutter et al., 2023). Briefly, raw gene count’s were normalized and transformed into log values using Variance Stabilizing Transformation (VST) as described previously (Love et al., 2014). Differential expression between groups and tissue were assessed using one-way ANOVA, and when significant effects were observed, Student’s t-tests were applied for pairwise comparisons. FDR correction was performed following Benjamini-Hochberg method (Benjamini et al., 1995).

### Pathway Enrichment and Protein Interaction Analysis

Differentially expressed (DE) genes identified in IMAT (p < 0.01) were analyzed using Ingenuity Pathway Analysis (IPA; QIAGEN Inc., https://www.qiagenbioinformatics.com/products/ingenuity-pathway-analysis). Canonical pathway enrichment analysis was performed using default settings, considering experimentally validated interactions. Significantly enriched pathways were ranked based on −log(p-value), with higher values indicating stronger statistical enrichment among DE genes. Pathways were subsequently curated to retain those with established relevance to adipocyte-related signaling, and the top enriched adipocyte-associated pathways were selected for downstream analysis and visualization.

EBF2 interaction partners were identified using the STRING database (version 11.5; https://string-db.org/), which integrates known and predicted protein–protein associations derived from experimental data, computational prediction methods, and public text collections (Szklarczyk et al., 2023). Network analysis was performed using default parameters, only direct interactions were considered. Identified interaction partners were used for downstream analysis and interpretation of EBF2-associated regulatory networks.

### Use of LLM in methods

A large language model (ChatGPT, OpenAI) was used to assist in generating portions of the R code for data visualization (including horizontal bar graphs, and correlation plots) and to support the classification of Ingenuity Pathway Analysis canonical pathways into biological themes for visualization. All outputs were reviewed, validated, and, where necessary, modified by the authors. The authors take full responsibility for the accuracy and integrity of the final content.

### Declaration of AI-assisted technologies in the writing process

We used ChatGPT (model: GPT-5.2) to improve readability and English language of the manuscript. After using this tool, the authors reviewed and edited the content as needed under strict human oversight and took full responsibility for the content of the publication.

## Data availability

Human bulk RNA-sequencing datasets are available from the authors upon reasonable request, subject to applicable data protection regulations.

The murine spatial RNA-seq data generated in this study have been deposited in ArrayExpress under accession number E-MTAB-17271.

## Author contributions

DL and SMH conceived the research question and designed and planned the study. RZT, MK, YZ, MW, SS, AF, KAD, BCB and MAP contributed to the acquisition of data. EP, RZT, MK, YZ, SS and DL analysed the data. DL and SMH interpreted the results and wrote the manuscript. All authors contributed to revision of the manuscript and approved the final manuscript prior to submission. PTP contributed expertise in mouse physiology and data interpretation and proofread the manuscript. DL and SMH are responsible for the integrity of the work as a whole.

## Acknowledgements

We acknowledge the technical support of Core Facility Core Facility Pathology and Tissue Analytics (CF-PTA), Core Facility Genomics (CF-GEN) and Core Facility Laboratory Animal Services, CF-LAS at Helmholtz Munich. We thank Dr. Inti Alberto de la Rosa Velazquez, Dr. Melisa Gómez, Mrs Ulrike Buchholz, Mrs Claudia-Mareike Pflüger for their assistance with spatial transcriptomics library sequencing, FASTQ file generation, skeletal muscle sectioning and placement on Visium slides, as well as antibody staining and microscopy. We thank Sebastian Cucuruz for support in animal handling, tissue isolation and histopathological imaging quantification of quadriceps sections. We thank Larissa Neuhauser for support in cell culture experiments and western blotting.

Open Access funding enabled and organized by Projekt DEAL. This work was supported by the Helmholtz Association; the German Research Council (DFG) grants HO 2286/3-1 (iMAGO, subproject P4) to SMH and the DFG grant 455422993 (iMAGO, subproject TP3, GZ: PL825/3-1) to MAP, both part of the Research Unit FOR 5298; the DFG grant 536691227 within the Research Unit FOR 5795-1 (HyperMet) to KAD and to DL; the DFG grant 545363987 (CoMeT) to DL; the European Research Council ERC-CoG Yoyo-LepReSens no. 101002247, to PTP.; the ADA grant 1-14-CE-05, the National Institutes of Health grant R01DK089170, the NIH General Clinical Research grant RR-00036 and the Colorado Nutrition Obesity Research Center grant P30DK048520 to BCB.

---

**Figure S1.**
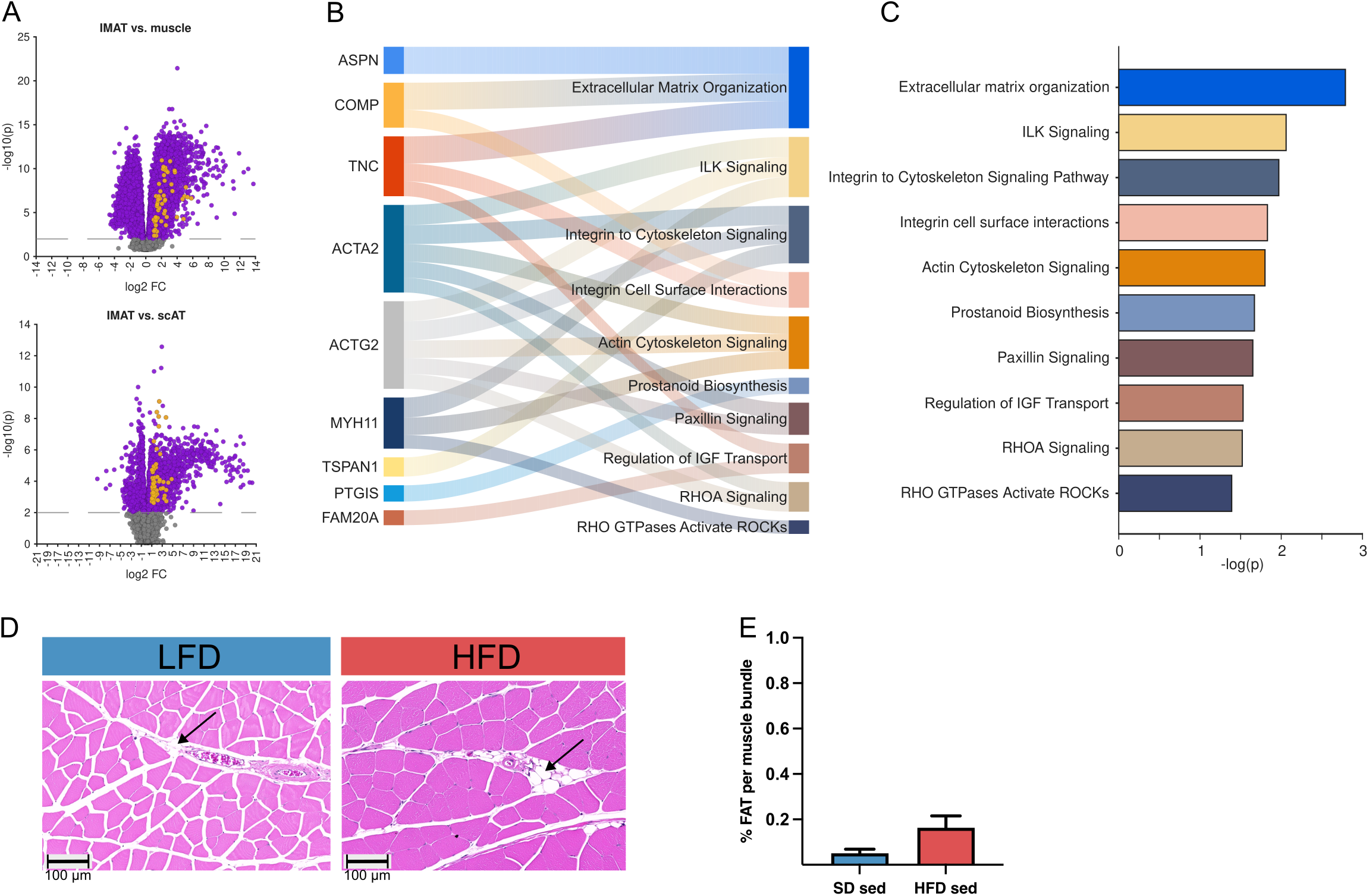
A) Volcano plots for all detected genes comparing expression between IMAT vs muscle (upper panel) and IMAT vs scFAT (lower panel). Genes selected as IMAT specific are highlighted in orange. B) Sankey plot of top adipocyte-related canonical signaling pathways identified by IPA and enriched in human IMAT-specific genes shown in the Fig. 1A heatmap, with corresponding gene-pathway associations C) Barplot showing enrichment -log10 p-values for selected adipocyte-related canonical signaling pathways. D) Representative histological sections of muscle quadriceps in high-fat-diet (HFD) and low-fat-diet (LFD) female mice. Intermuscular adipocytes are indicated by arrows. Scale 100um. E) Quantitative digital imaging analysis of histological sections in female mice (n = 7-8) after 84 days of HFD and LFD feeding. Data are presented as mean ± SEM.

**Figure S2.**
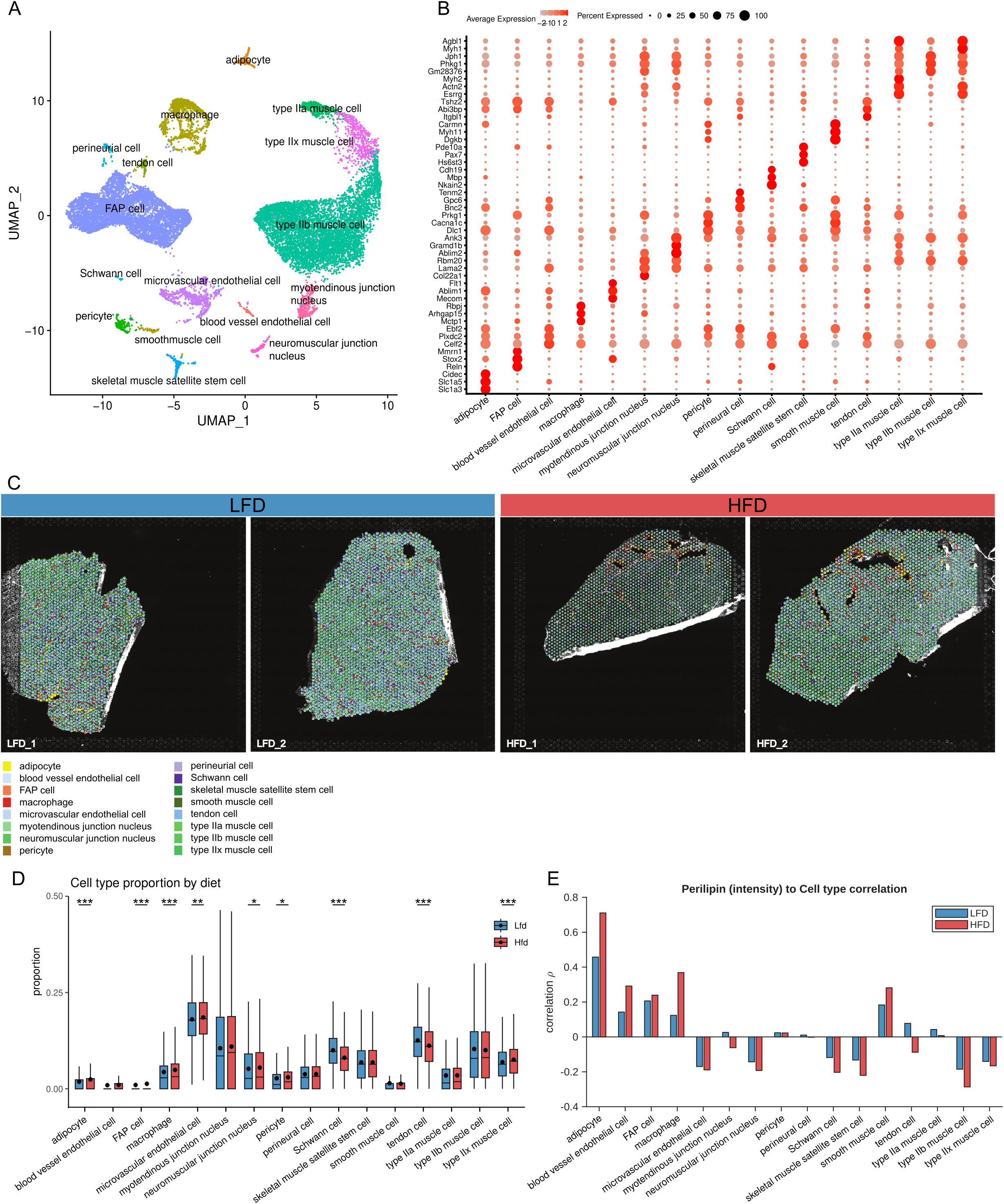
A) UMAP of the murine snRNA-seq reference dataset, colored by cell-type annotation. B) Dot plot displaying expression levels of the top three marker genes, selected based on mean AUC, for each reference cell type across all identified cell types. C) Spatial pie chart plot showing spotwise cell-type distributions in LFD and HFD fed male C57BL/6 mice, shown on Visium 10x Genomics slides (n = 2). D) Boxplots showing the distribution of inferred cell-type proportions in spatial transcriptomics spots from LFD and HFD fed male mice on Visium 10x Genomics slides (n = 2). E) Bar plot of correlation coefficients comparing spotwise perilipin intensity with cell type content for LFD and HFD fed male mice, acquired on Visium 10x Genomics slides (n = 2).

**Figure S3.**
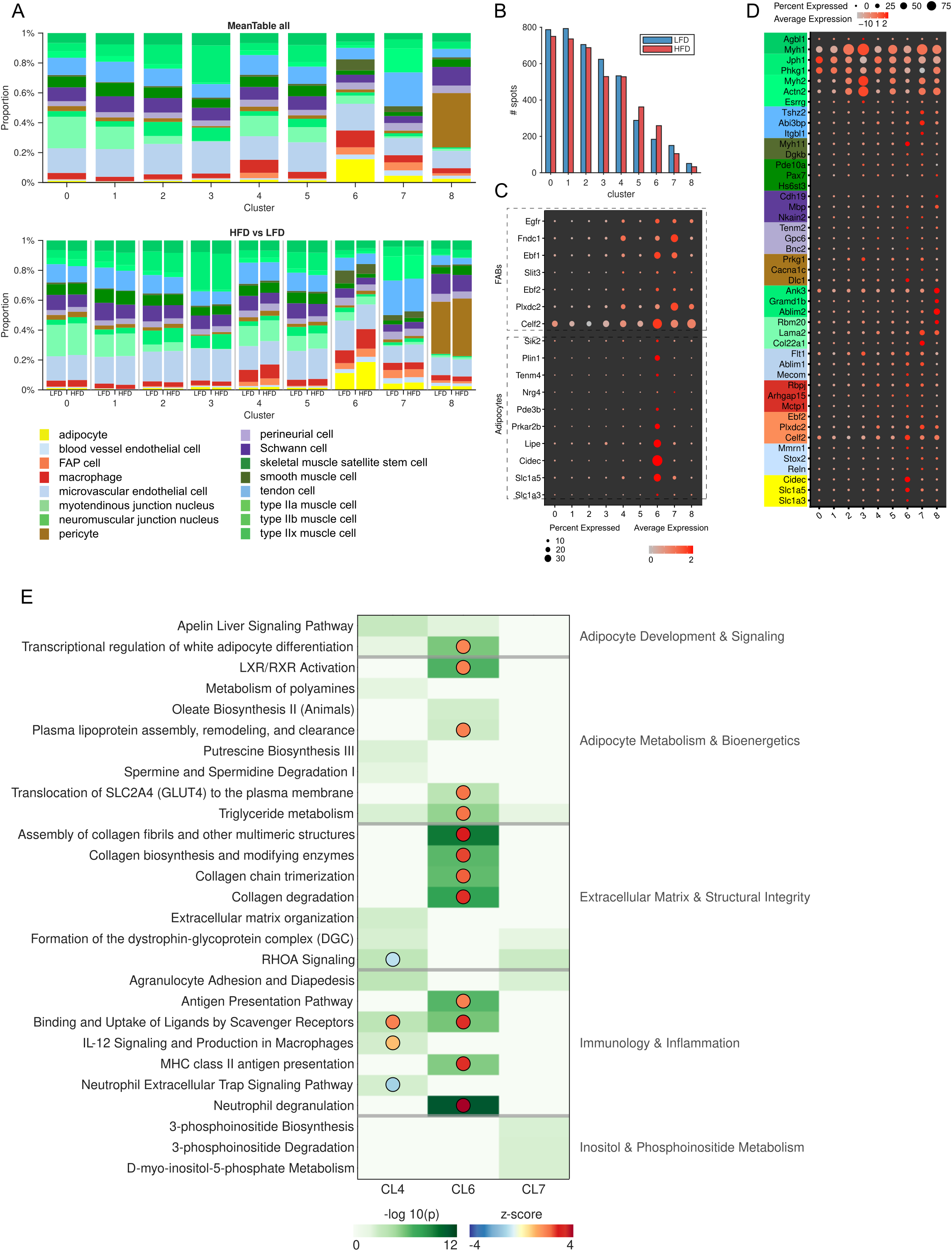
A) Proportional cell type distributions over all 9 identified clusters averaged over all spots and samples (top) and diet specific (bottom). Stacked barplots refer to the mean cell type proportions over all spots for each cluster. B) Barplot of total spots mapped to the 9 clusters between high-fat diet (HFD) and low-fat diet (LFD) male mice. C) Dot plot of adipocyte and FAP marker genes from the reference dataset across all nine spatial clusters. D) Dot plot displaying expression of the top three marker genes for each reference cell type across all nine clusters. E) Selected Ingenuity Pathway Analysis (IPA) results for genes differentially regulated in HFD compared to LFD male mice, identified across spatial transcriptomic clusters 4, 6, and 7. Color intensity reflects enrichment magnitude (p-value), and pathways are grouped by biological theme to highlight the functional landscape driven by HFD-associated gene expression. Overlaid dots indicate the IPA activation z-score where calculable — pathways lacking a dot either contained fewer than four analysis-ready molecules from the input dataset or showed insufficient directional concordance with curated pathway relationships, precluding z-score estimation.

**Figure S4.**
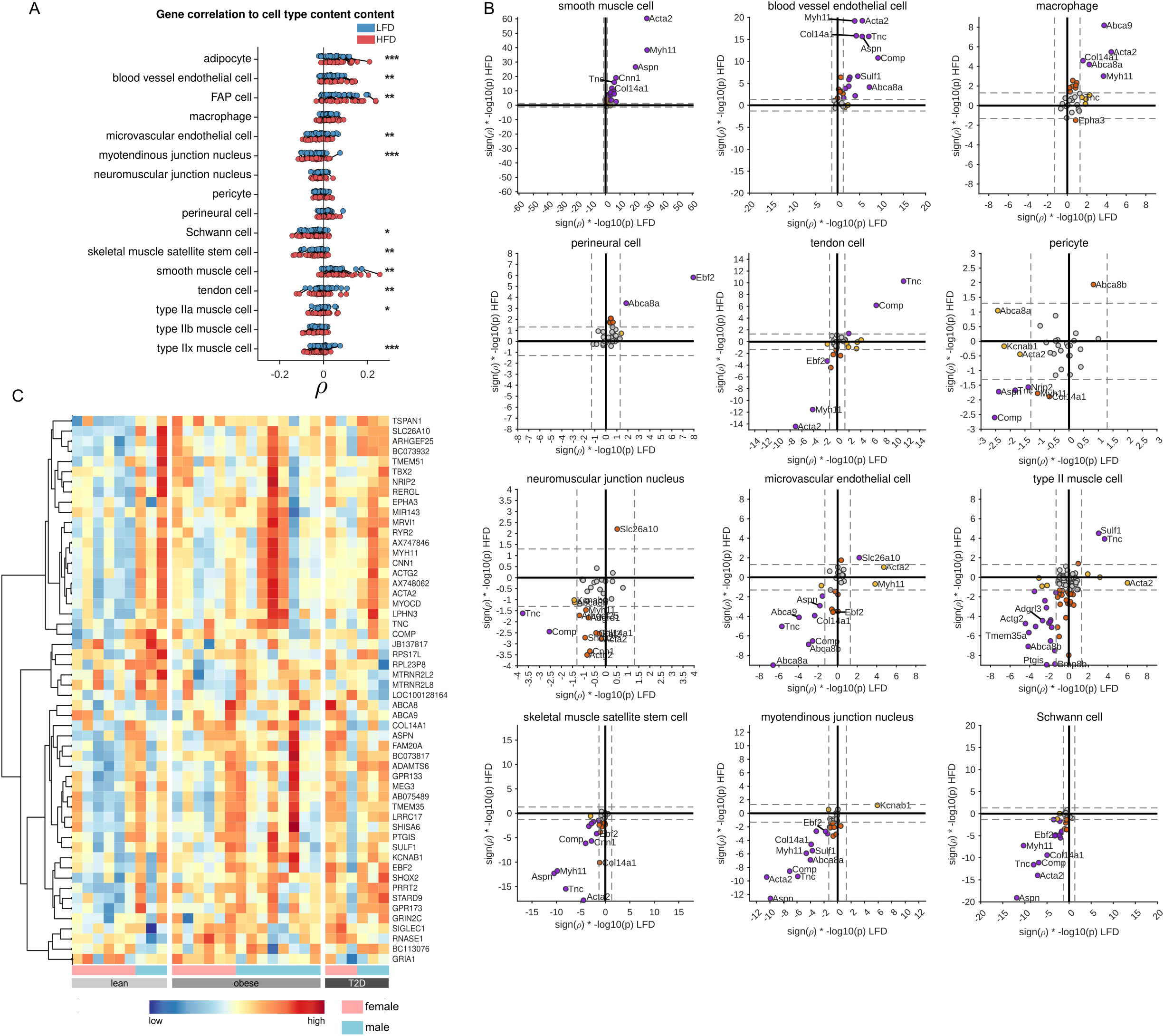
A) Spearman rank correlation of human IMAT gene signature comparing spot wise expression and cell type proportions in murine skeletal muscle from low-fat diet (LFD) and high-fat diet (HFD) fed male C57BL/6 mice (n = 2). B) Scatterplot of directed p-values (p * sign (rho)) comparing IMAT specific correlation between spot wise human IMAT gene expression and cell type proportion for all identifued cell types (except adipocytes and FAPs) in LFD fed (x-axis) and HFD (y-axis) fed male mice. Purple dots denote genes significantly correlated in both directions; orange dots denote genes correlated in x-dimension (LFD) only and red in y-dimension (HFD) only. C) Heatmap of human IMAT gene signature in female and male lean, obese and type 2 diabetes (T2D) participants.

**Figure S5.**
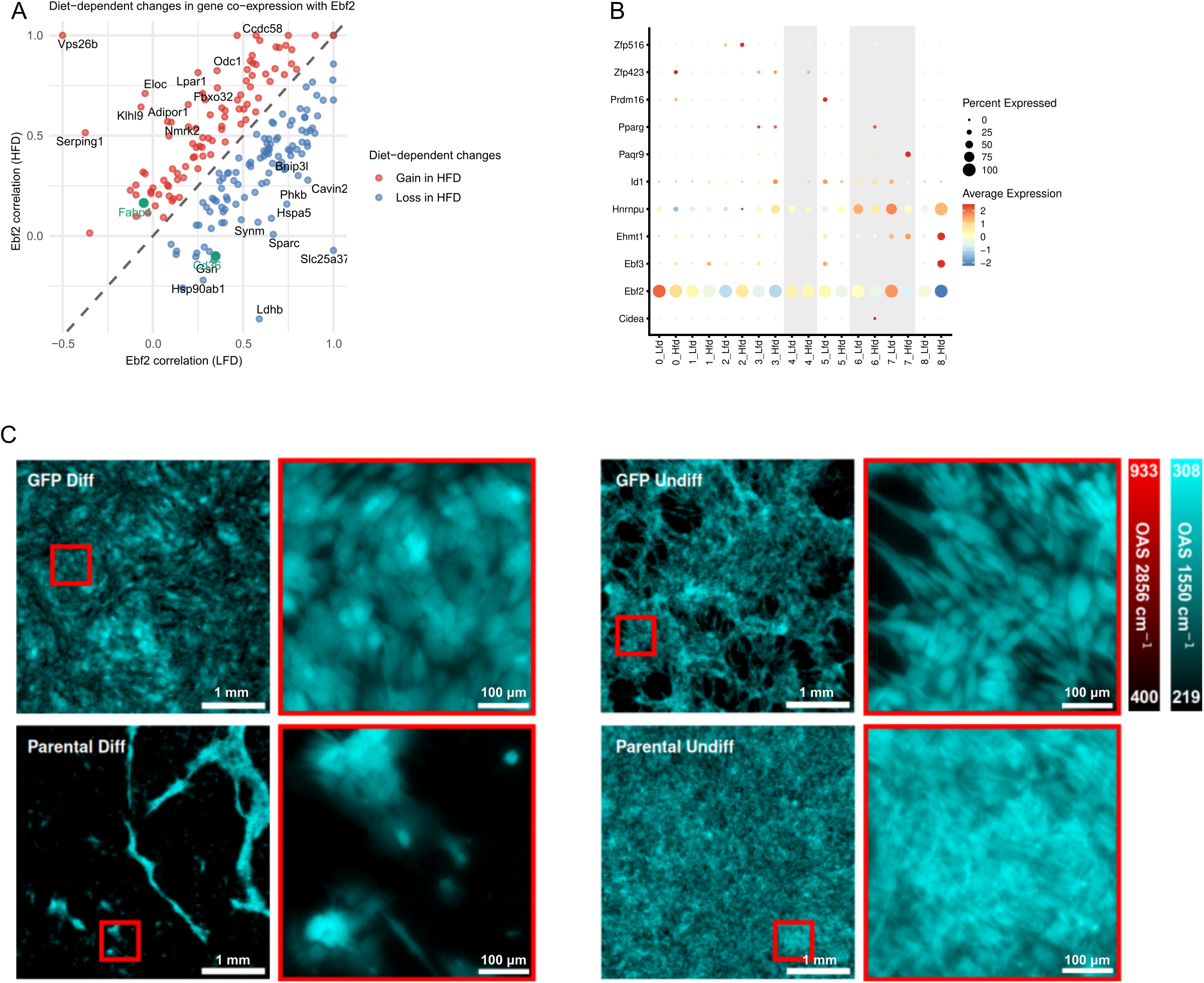
A) Spatial transcriptomic analysis of genes co-localizing with Ebf2 in C56/Bl6 male mice. Each point represents a gene co-expressed with Ebf2. The x- and y-axes show Spearman correlation coefficients between Ebf2 and the indicated gene in low-fat diet (LFD) and high-fat diet (HFD) barcodes, respectively, calculated using pairwise complete observations. The dashed diagonal indicates unchanged correlation across diets; points above or below the diagonal represent genes with increased or decreased correlation under HFD. Selected genes with the strongest diet-dependent changes are labeled. Ebf2 regulated genes Fabp4 and Cd36 are highlighted in green. B) Dot plot showing expression of STRING-identified EBF2 interaction genes across nine spatial transcriptomic clusters from LFD and HFD fed male C57BL/6 mice (n = 2). C) Human skeletal muscle cell (HSkMC) adipogenic differentiation (Diff.) and non-differentiation (Undiff.) comparative analysis by mid-infrared optoacoustic microscopy (MiROM). OA micrograph of lentiviral GFP gene overexpression in HSkMC (upper panel) and parental (wt) HSkMC (lower panel), Big FOV (4 mm × 4 mm) is on the left side, and small FOV (500 μm×500 μm) is on the right side in two channels: red (at 2856 cm^-1^; CH_2_ band - symmetric CH_2_ stretching) and cyan (at 1550 cm^-1^; Amide II band – N-H bending/ C-N stretching).

